# Environmental and social impact assessment in healthcare business planning: a national survey of NHS trusts in England

**DOI:** 10.1101/2024.11.13.24317090

**Authors:** Haruna Takahashi, Søren Kudsk-Iversen

## Abstract

**Background:** Decision-making in healthcare should include considerations for its environmental and social consequences as well as clinical and financial. Business cases represent projects which are likely to have particularly high impact, in which performing an Environmental and Social Impact Assessment may reduce negative effects. The practice of assessing these impacts is not mandatory, therefore our aim was to determine to what extent environmental and social considerations are incorporated into business planning across NHS trusts in England. We also set out to identify examples of good practice, common challenges and possible solutions to overcome these.

**Methods:** We conducted a cross-sectional survey across trusts in NHS England to capture current practice. This was carried out online between April and July 2024.

**Results:** Ninety seven out of 210 trusts (46%) responded to the survey. Thirty two currently assess the environmental and/or social impact of their business cases and 25 are in the process of doing so, illustrating a growing practice. There is significant variation between trusts in terms of content, format and process of the assessment.

**Discussion:** This was the first study to portray not only whether and how environmental and social impacts are assessed in NHS business planning, but also the barriers and potential solutions in embedding this assessment. The variation in practice signifies heterogeneity of NHS trusts and there is unlikely to be a single tool that would suit every organisation. An impact assessment process with a clear purpose, that reflects organisational values, and is supported by multidisciplinary expertise is likely to be more meaningful. We include an Environmental and Social Impact Assessment tool for our trust, which has been developed based on the learning points from this study and modified to our trust’s needs, as an example.

## Introduction

Decision-making in healthcare is influenced by many factors including clinical and financial. Projects that require submission of a business case proposal for financial justification and organisational approval represent particularly high-cost decisions that are likely to have significant environmental and social consequences. As NHS Trusts in England reach their first interim target in 2028-32 of an acute carbon footprint 80% less compared to 1990 levels, they need to assess these high value projects at the conception stage, to ensure their true impacts are considered and improved upon where required. In the UK, sustainability impact assessments are an established practice in many sectors such as business and infrastructure planning [1, 2]. However, it is not routine in healthcare despite its large climate and social footprint [3]. As well as the legal net zero targets of 2040 and 2045 for the NHS, there is a strong case for reducing the negative impact of healthcare-related activities which, ironically, is leading to a health crisis and health inequalities [4, 5].

This results in an increased strain on the health service leading to a downward spiral. Healthcare systems can therefore limit the impact on the planet, people and itself, by mitigating the impact of its own activities. This aligns with many aspects of NHS England’s long term plan, which includes promoting population health and tackling health inequalities [6].

NHS England recognised the need for greener business cases in their document *Delivering a ‘Net Zero’ National Health Service* in 2022. Two years on, healthcare organisations are still not required to consider its environmental or social impact, with the exception of procurement tenders [4, 7, 8]. Nevertheless, NHS England’s *Greening the business case* provides some guidance in the realm of estates [9]. This guidance highlights some of the arguments for greener capital investment: improving population health, reducing health inequalities, creating local jobs in the low carbon economy, exerting influence as an anchor institution, adapting to adverse effects of climate change, reducing cost, and improving patient outcomes. These should be considered more broadly in all investment types. Beyond this document there is no directive from NHS England for organisations to assess the environmental or social impact of their investments, leaving individual trusts to decide whether and how to incorporate this into their business planning. Furthermore, there is little information available on current practice, which is crucial to ascertaining the next steps towards embedding this into routine practice nationwide.

The aim of this survey was primarily to identify to what extent environmental and social considerations are incorporated into business planning across NHS trusts in England. Additionally, we explored examples of good practice and challenges identified, to enable organisations, including our own, embed this process more effectively as well as incite national guidance.

## Methods

We conducted a cross-sectional survey via email using an nhs.net email account, between April and July 2024. The survey protocol was assessed by the Joint Research Office at Oxford University Hospitals NHS Foundation Trust and deemed exempt from ethics review. The term ‘environmental and social impact assessment (ESIA)’, rather than ‘sustainability impact assessment’, will be used throughout to differentiate from organisational sustainability, and to reflect the close interlink between planetary health and social value. The survey is reported according to the Checklist for Reporting Of Survey Studies (CROSS) reporting guideline [10].

We included all existing NHS trusts and foundation trusts in England, derived from the NHS England provider directory (accessed in April 2024) [11]. We identified a contact in each trust that was most likely to be able to provide the required information using a stepwise approach:

1. If available on trust websites, we contacted the trust sustainability lead (or equivalent)
2. If not available or no response within two weeks, we contacted the trust sustainability team email address
3. If not available or no response within two weeks, we contacted the generic trust email address
4. If no response within two weeks, and none from any of the previous attempts, we sent a follow-up email to the first contact approached
5. If no response within two weeks after follow-up, the trust was considered a non-responder

Once an appropriate contact was identified, they were asked if their trust business cases included any information on environmental and/or social impact. If yes, they were asked a further set of questions regarding the content, format and use of their environmental and social impact assessment (appendix 1). They were also requested to share their impact assessment questions or tools. We did not pretest the questionnaire.

Responses were anonymised by allocating a random number to each trust at the time of first contact and collated on an Excel spreadsheet. The key was accessible to the lead investigator (HT) only, in case further correspondence was required to clarify answers to the survey. Once the data collection phase had been completed the key was destroyed, leaving only the anonymous survey data.

Quantitative data was presented in a descriptive manner using total count and percentages with no statistical hypotheses formulated or tested. Qualitative data was summarised thematically. No sensitivity analysis was performed.

## Results

Number of trusts at each stage of the data collection phase is illustrated in Fig 1. Out of 210 trusts contacted, 97 responded to the survey giving a response rate of 46%. Of these, 57 had either an ESIA in place (n=32) or was in progress (n=25). Five trusts required a Freedom of Information request to provide contact details for staff members. Two trusts had a shared sustainability lead and team.

**Fig 1.**
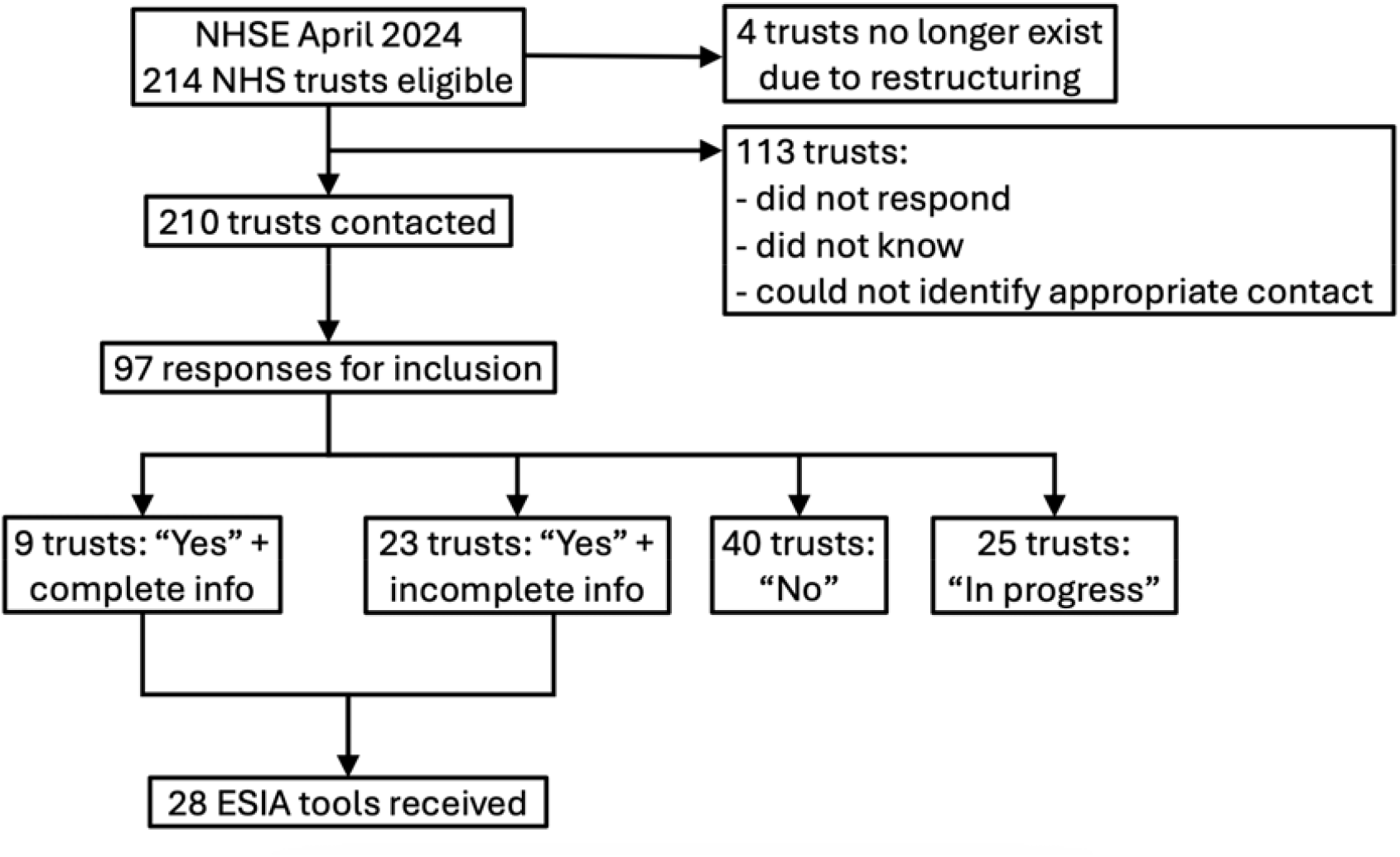
Number of trusts at each stage of data collection.

The characteristics of respondents and those using ESIA were broadly similar to the overall eligible population (table 1). Contacts with an official role in sustainability represented the majority, at 72% of all respondents and 84% of those in trusts that that currently practise ESIA.

**Table 1.** Characteristics of trusts and contacts included in the survey. * Two trusts had a shared sustainability team

|  | Trusts<br>contacted<br>(n=210) | Trusts that<br>responded<br>(n=97) | Trusts that<br>practise ESIA<br>(n=32) |
| --- | --- | --- | --- |
| **Trust type** |  |  |  |
| NHS trust | 67 (32%) | 32 (33%) | 13 (41%) |
| NHS foundation trust | 143 (68%) | 65 (67%) | 19 (59%) |
| **Service category** |  |  |  |
| Acute | 113 (54%) | 55 (57%) | 19 (59%) |
| Mental health | 24 (11%) | 12 (12%) | 5 (16%) |
| Community health | 17 (8%) | 4 (4%) | 1 (3%) |
| Ambulance service | 10 (5%) | 6 (6%) | 2 (6%) |
| Mixed | 46 (22%) | 20 (21%) | 5 (16%) |
| **Contact's role type** |  |  |  |
| Sustainability |  | 70 (72%)* | 27 (84%) |
| Finance |  | 7 (7%) | 1 (3%) |
| Sustainability and finance |  | 2 (2%) | 0 (0%) |
| Estates |  | 7 (7%) | 1 (3%) |
| Strategy |  | 6 (6%) | 0 (0%) |
| Other |  | 5 (5%) | 3 (9%) |

Table 2 summarises the findings of additional questions regarding ESIA. Better response rate was seen from trusts who provided the impact assessment tools. Three trusts reported to have based their ESIA on sustainability models; these were Centre for Sustainable Healthcare Sustainability in Quality Improvement framework (two trusts), and Triple Bottom Line and Doughnut Economics (one trust), [12, 13, 14]. Five were copied or adapted from other trusts and two were based on the trusts’ own sustainability strategies. Two trusts reported their ESIA tools were standardised throughout their integrated care systems. In 12 trusts their ESIAs were original.

**Table 2.**
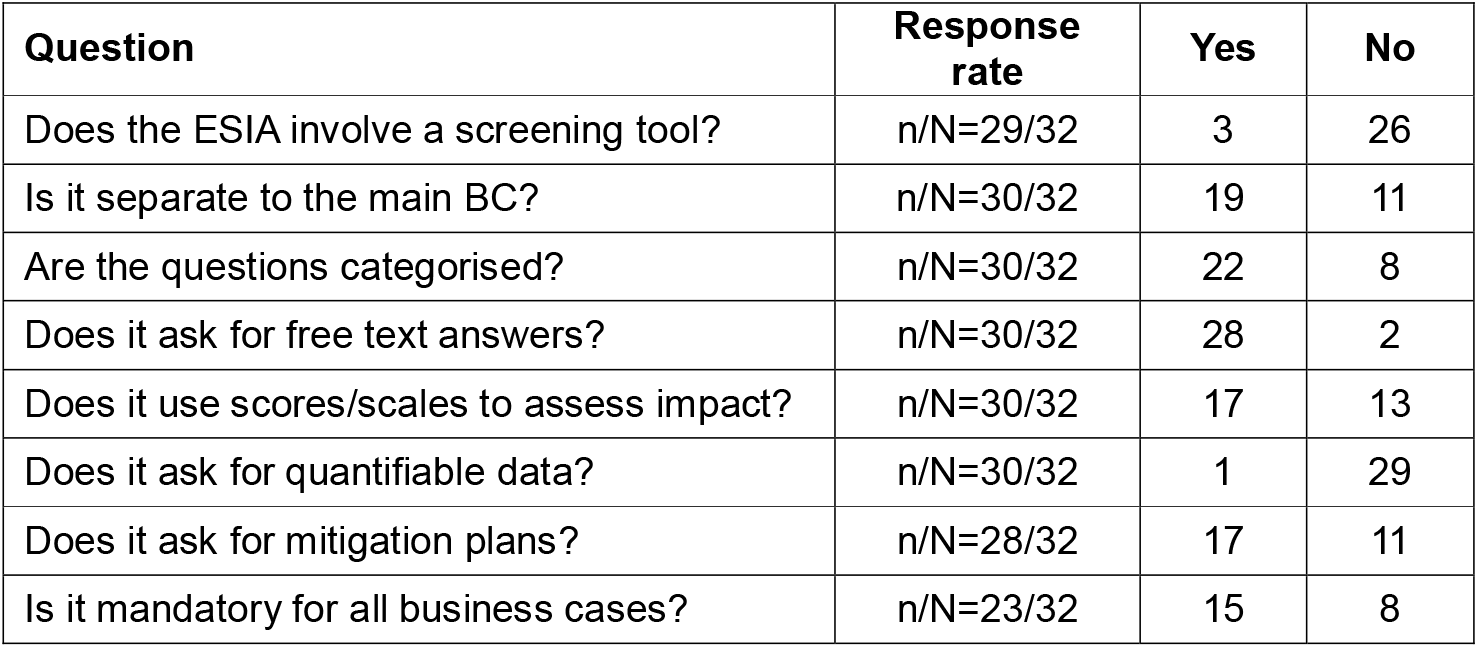
Findings related to content, format and process of ESIA use. ESIA = environmental and social value impact assessment, BC = business case.

Screening questions included in three of the ESIAs were used for different purposes: to determine whether the full ESIA should be completed, to determine which sections of the ESIA should be completed, and to determine whether the sustainability manager should be contacted.

Twenty three out of 32 trusts provided information on whether ESIA was required for business cases. This showed that ESIA was compulsory for all business cases in 15 trusts, six mandated it for business cases meeting certain criteria such as a cost threshold or investment type, and two trusts had their ESIAs as optional components of their business cases. Twenty one trusts provided information regarding the review process for completed ESIAs. They were routinely reviewed in 14 trusts, 12 of which by the sustainability lead or team. The review led to formal approval by either the sustainability or the business planning team in five trusts, whereas comments and/or advice alone was offered in eight. Five trusts did not review their ESIAs. Following business case approval, four trusts had an established system to follow up some or all of their ESIAs separately to business case follow up.

Four were unsure if the introduction of an ESIA had changed the business planning practice in their trust, while two felt it made no difference, and nine said it was too early in the process to tell. However, four respondents felt it had made a difference, although without any metrics in place to measure this. These four respondents all had sustainability roles in acute trusts, three of whom were heads of sustainability. The ESIA was compulsory in all four organisations, although in one trust this only applied to capital investment. Their ESIAs all contained questions categorised into sustainability and social value themes and asked for a combination of free text and impact scores. One ESIA involved screening to identify net increase in carbon footprint which in turn directed the author to contact the sustainability manager, while the remaining three ESIAs asked for mitigation plans. In all four the completed ESIAs were reviewed by the sustainability team who offered comments and advice but was not required to give formal approval. Only two consistently provided followed up.

Table 3 summarises barriers encountered during implementation and potential solutions offered by the respondents which have been successful in some trusts. Most barriers were related to the process, rather than content, of ESIAs.

**Table 3.** Barriers and successes experienced by trusts when embedding, maintaining and improving the environmental and social impact assessment. ESIA = environmental and social impact assessment.

| **Barriers** | **Potential solutions** |
| --- | --- |
| Creating a tool that is objective | • Balancing quantitative data (objective and useful) and qualitative data (more easily obtained) |
| Lack of sustainability team workload capacity | • Filtering business cases or aspects within cases with high impact using screening questions or impact scores<br>• Prioritising business cases with higher financial cost<br>• ESIA tools with built-in guidance function (such as instructions, automated calculations, and links to resources) |
| Multiple business planning pathways, forms and committees | • Close partnership with business planning group(s) |
| Additional workload for business case authors and business planning team | • Embedded screening tool to ensure amount of information requested is proportional to the size of the business case |
| ESIAs not completed or sent to sustainability team | • Sustainability team/lead included in business case review panel<br>• Sustainability lead approval required for business case to go ahead |
| Lack of engagement from stakeholders | • Better understanding of rationale for the impact assessment through education, training and discussion<br>• Representatives from teams such as sustainability, estates and strategy involved in business planning |
| Lack of sustainability expertise in business case authors | • Including questions that can be answered by business case authors (with or without input from other resources)<br>• Links to helpful resources included in ESIA form<br>• Sustainability included in business case training<br>• Provision of resources to help impact assessment such as carbon measurement tools<br>• ESIA incorporated into quality improvement programmes, which helps increase awareness and knowledge in departments before business cases are initiated |
| Deciding what to do with ESIA outcome and who is accountable | • Clear purpose for the ESIA<br>• Agreement between sustainability and business planning teams on the use of ESIA |

## Discussion

This is the first survey in the NHS in England investigating how sustainability is incorporated into business planning at trust level, offering vital information to help organisations embed the practice successfully. It has demonstrated that the proportion of trusts currently assessing the environmental and social impact of their business cases is small but likely growing. Fifty seven out of 97 respondents (59%) either have an impact assessment in place or are in the process of implementing one, and this number is likely to increase as we approach the first interim net-zero target in 2028-32. This is encouraging and may help motivate organisations currently without ESIAs.

A wide variation in practice was observed in all aspects of impact assessment, namely content, format and process. Whilst this may be attributed to lack of guidance and standardised approach, it may also reflect the heterogeneity of NHS trusts in England, including size, management structure, priorities, business planning pathways, clinical activities, and sustainability team size and structure. This is evident in barriers to implementation identified, where trusts reported different challenges. In view of the multiple complex factors that need to be taken into consideration, there is unlikely to be a single assessment tool or process that is suitable for all NHS organisations; in other words, there is no ‘one size fits all’. Trusts must therefore take these factors into account when building a new ESIA tool or implementing its use. At a national level, the complexities are beyond simply publishing an ESIA tool and mandating its use.

While trusts must find a process that suits them, we found common themes in those that appeared to be successful. Characteristics shared by trusts that reported a positive change reinforce the importance of a robust ESIA process (table 4).

Respondents with sustainability roles represented a higher proportion in trusts that had ESIAs. They were also the only ones to report possible beneficial effects of ESIAs in business cases in their trusts. The presence of a dedicated sustainability team represents expertise and workload capacity, and it may also reflect organisations that prioritise sustainability. Although all NHS trusts in England must have a board-level net-zero lead, the presence, size and activity of sustainability teams vary [4]. Having said this, the involvement of a sustainability team should complement, not detract from, other stakeholder engagement, as partnership seems to be crucial in the meaningful utilisation of ESIA. To provide the reader with an example of a possible ESIA building on these themes, we include the current draft ESIA for Oxford University Hospitals NHS Foundation Trust (appendix 2). It was created based on the findings of this survey and has been developed by HT with assistance from a wider multidisciplinary team, including operational managers and the trust leads for sustainability and anchor approach, to list but a few. Importantly it is unique to the Trust and its current set up, and is an evolving process. We intend to review its roll-out during 2025 with a plan to disseminate our findings.

Many trusts found it challenging to ensure their ESIAs had any influence on their business cases. Decision-making in the NHS is largely a balance between clinical need and financial constraint, despite it being considered a service, not a business [15, 16]. As a service, its interests should lie not only in health gain and financial impact, but also its environmental and social values [17]. The responsibility of the NHS extends beyond immediate health needs of its current patients to include longer term wellbeing of the whole population [18]. Business cases within NHS organisations should reflect this overall aim, and their worthiness should be assessed as such [19]. Rather than evaluating “value for money” where ‘value’ equates to short term clinical outcomes, we should be evaluating “value for cost” where’ value’ is redefined to reflect population health and social value, and ‘cost’ includes other finite resources such as the natural environment [17, 19]. It would seem sensible therefore to first redefine the purpose of business case evaluation, instead of merely adding an extra piece of mandatory paperwork. In fact, excessive collection of data can lead to ethical, legal and practical problems, such as information silos representing wasted data and resources [20, 21]. Redefining the terms ‘value’ and ‘cost’ may be context-dependent, again owing to the heterogeneity of NHS organisations across England. For example, a trust may decide to focus on simply reducing the carbon emission of each business case, while another may decide to include broader issues such as air pollution and local health promotion.

This in turn would reflect trust aims, strategies and regional characteristics. Furthermore, each trust has a different management structure and methods of sharing data between departments that will influence how such an impact assessment can be executed.

Measuring impact was another challenge identified by the survey. The effect of this is two-fold; first, it complicates the impact assessment itself. For instance, carbon footprint is one way to measure environmental impact but other effects such as biodiversity loss are harder to quantify, and there is no standardised method of measuring social value [15]. Not only does this pose a risk of missing some of the impacts, but it also makes it difficult to determine the size of the impact. Second, it is hard to ascertain the effect of any impact assessment process as there are no figures akin to financial savings. Despite this there are benefits beyond those that can be measured, and solely focusing on measuring impact can be counterproductive [15, 22]. For instance, an assessment tool can provide a teachable moment to educate and change behaviour [23]. We suggest trusts use the environmental and social impact assessment as an opportunity to embed sustainable values into every decision-making.

There were some limitations to this study. Firstly, whilst all existing trusts had been contacted, the response rate was not 100%. It is possible that trusts that responded were more likely to have an impact assessment in place, due to confounding factors such as the presence of an active sustainability team and an inclination to take part in the survey if the answer is positive. This may have led to an overestimate of trusts practising ESIAs. Similarly, the missing data on further questions may be due to the same bias where those with more to contribute were more likely to answer. Secondly, although we tried to identify the person with the most knowledge in this topic in each trust as our contact, some trusts could not identify an appropriate person. In those that did, there was variability in the respondent’s role and involvement in business cases. This may have resulted in incomplete or incorrect information. Thirdly, some questions permitted subjective answers based on the respondent’s opinion.

Interpretation of the questions may have varied between individuals despite efforts to make them as unambiguous as possible. For example, a “change in sustainable practice in business planning” could refer to ESIA completion rate, content, increased awareness or influence on business case. Fourthly, there may have been reporting and recall bias as the survey relied on answers by contacts rather than collecting recorded data. Finally, analysis of trusts with seemingly beneficial impact assessment process needs to be interpreted carefully, due to the small number and assumptions made. Nevertheless, evaluation of good examples is helpful in advancing individual and national performance.

## Conclusion

The assessment of environmental and social impact of NHS business cases in England is an evolving practice. In order to meet our legal and moral obligations of improving sustainability and adding social value through healthcare, every decision, including those requiring business cases, must involve an evaluation of its impacts. National guidance would help accelerate and standardise practice, although heterogeneity within the NHS means every trust needs to adjust the process to suit them. Whilst sustainability team input is crucial, business case authors and other stakeholders should be educated and encouraged to participate, as all NHS staff have a role in creating a sustainable healthcare system [4].

## Contributors

Haruna Takahashi: Conceptualisation, investigation, data analysis, writing – original draft preparation. Søren Kudsk-Iversen: Conceptualisation, supervision, writing – reviewing and editing.

## Data Availability

All data produced in the present study are available upon reasonable request to the authors

## Data availability statement

Data are available upon reasonable request.

## Patient consent for publication

Not applicable.

## Ethics approval

Not applicable.

## Acknowledgements

This study was performed as part of Specialty Training in Anaesthetics with the support of the Royal College of Anaesthetists, UK, although the views and opinions expressed in this paper are those of the authors and responsibility for the content remains exclusively with the authors. We thank Dr Ruth Webster, Training Programme Director, and Dr Amy Swinson, College Tutor, Thames Valley School of Anaesthesia, for their support in incorporating this into the anaesthetic training programme.

We appreciate the input from Kate Townsend, Greener NHS, who directed us to useful resources and contacts.

We are grateful to all survey participants for their time and insight into their work.

