## Appendix 1 for "Environmental and social impact assessment in healthcare business planning: a national survey of NHS trusts in England"

Appendix 1 – Questions included in the survey sent to trusts who said they practised environmental and social impact assessment (ESIA)

| **Questions about content and format of ESIA** | **Questions about the process of using the ESIA** |
|---|---|
| Was it based on a particular model or adapted from other tools? | Is it mandatory to complete? |
| Does it involve a screening tool? | Who reviews the completed ESIA? |
| Is it separate to the main business case proposal form? | Does it require formal approval? |
| Are the questions categorised into environmental and/or social themes? | Is it followed up any time after business case approval? |
| In what format are the answers? (Free text, scores/scales, or quantifiable data) | Has the introduction of the ESIA changed business planning practice, and if so has this been measured? |
| Does it ask for mitigation plans? | What were the barriers and challenges when embedding this into practice, if any? |
