## Appendix 2 for "Environmental and social impact assessment in healthcare business planning: a national survey of NHS trusts in England"

Appendix 2 – A draft Environmental and Social Impact Assessment tool for business cases in Oxford University Hospitals NHS Foundation Trust

Oxford University Hospitals NHS Foundation Trust

### Environmental and social impact assessment

Business case title: ______________________ Business case authors: ______________________

| | Category | OUH strategic theme | Short-term impact | Long-term impact | Actions to mitigate negative impacts and enhance positive impacts (SMART objectives) | Considerations | Resources |
|---|---|---|---|---|---|---|---|
| Environmental | ENERGY | | Neutral | Positive | | • Fossil fuels, electricity and battery usage<br>• Energy efficient estates<br>• Renewable energy | Greening the business case<br>Delivering a net-zero NHS<br>xxxxx@xxxxx (Head of Sustainability) |
| | WATER | | Negative | Neutral | | • Water consumption: low water efficiency equipment, leaks, landscape watering, kitchens/bathrooms/sinks<br>• Substance disposal into water | xxxxx@xxxxx (Head of Sustainability) |
| | WASTE | | Choose an item. | Choose an item. | | • Amount and type of waste generated<br>• Waste segregation and recycling | xxxxx@xxxxx (Waste management) |
| | TRAVEL | | Choose an item. | Choose an item. | | • Patient travel: care closer to home, fewer/joint appointments<br>• Business travel, NHS fleet<br>• Staff commute | Delivering a net-zero NHS<br>xxxxx@xxxxx (Green travel and transport manager) |
| | POLLUTION & BIODIVERSITY | | Choose an item. | Choose an item. | | • Air and water pollution<br>• Indoor air pollution<br>• Green spaces | Health matters: air pollution<br>Improving access to green space<br>xxxxx@xxxxx (Head of Sustainability) |
| | DIGITAL TRANSFORMATION | | Choose an item. | Choose an item. | | • Using virtual platforms to reduce paper, travel, energy and time<br>• Sustainable use of digital devices<br>• Digital patient platforms, digital inclusion | xxxxx@xxxxx (Digital and information) |
| | PROCUREMENT | | Choose an item. | Choose an item. | | • Whole life costs<br>• Local, ethical, sustainable suppliers<br>• Greener catering | Note: social value evaluation included in procurement tenders<br>Delivering a net-zero NHS<br>Ext XXXXX (Procurement) |
| Social | HEALTH PROMOTION | | Choose an item. | Choose an item. | | • Disease prevention and health promotion<br>• Promoting independence<br>• Reducing avoidable hospital/residential care admissions | OUH Here for Health<br>xxxxx@xxxxx (Health promotion) |
| | COVID-19 RECOVERY | | Choose an item. | Choose an item. | | • Supporting individuals affected by Covid-19<br>• Supporting businesses affected by Covid-19<br>• Supporting clinical services affected by Covid-19 | |
| | TACKLING ECONOMIC INEQUALITY | | Choose an item. | Choose an item. | | • Creating new jobs and skills<br>• Increasing suppy chain diversity, resilience and capacity<br>• Supporting small and local businesses | Anchor steering group<br>xxxxx@xxxxx (Workforce) |
| | WELLBEING | | Choose an item. | Choose an item. | | • 6 domains of wellbeing: emotional & phychological, physical, social, financial, occupational & intellectual, environmental<br>• Wellbeing champion | OUH wellbeing<br>xxxxx@xxxxx (Wellbeing) |
| | EQUAL OPPORTUNITIES | | Please complete separate Equality Impact Assessment | | | | |

| | | |
|---|---|---|
| Date reviewed:<br>Action plan: | Reviewed by: | Follow up: Y/N<br>Date of planned follow up: |

Created by Haruna Takahashi

*Where a cell asks to "Choose an item", the business case author will select from "Positive", "Neutral" or "Negative" impact. Energy and water categories illustrate this as examples.
